# External Validation of the Predicting Asthma Risk in Children (PARC) tool in a clinical cohort

**DOI:** 10.1101/2022.03.28.22273062

**Authors:** Daria Olena Berger, Eva S L Pedersen, Maria Christina Mallet, Carmen C M de Jong, Jakob Usemann, Nicolas Regamey, Ben D Spycher, Cristina Ardura-Garcia, Claudia E Kuehni, the SPAC Study Team

## Abstract

**Rationale:** The Predicting Asthma Risk in Children (PARC) tool uses questionnaire-based respiratory symptoms collected from preschool children to predict their risk of asthma 5 years later. The tool was originally developed and externally validated in population-based settings and has not yet been validated in a clinical setting.

**Objective:** To externally validate the PARC tool in children seen in paediatric pulmonology clinics.

**Methods:** The Swiss Paediatric Airway Cohort (SPAC) is a prospective study of children seen in respiratory outpatient clinics across Switzerland. This analysis included children seen at ages 1-6 years for cough or wheeze at baseline and who completed the follow-up questionnaire 2 years later. The outcome was defined as current wheeze plus use of asthma medication. In sensitivity analyses, we explored effects of varied inclusion criteria and outcomes. We assessed performance by describing sensitivity, specificity, negative and positive predictive value (NPV, PPV), area under the curve (AUC), scaled Brier’s score and Nagelkerke’s R^2^ scores and compared performance in SPAC to that achieved in the original population, the Leicester Respiratory Cohort (LRC).

**Results:** Among the 346 children included, 125 (36%) reported the outcome after 2 years. At a PARC score cut-off of 4, sensitivity was higher (95% vs 79%) but specificity lower (14% vs 57%) in SPAC compared to LRC. NPV was comparable (0.84 vs. 0.87) as was PPV (0.37 vs.0.42). Discrimination was lower in SPAC (AUC of 0.71 vs 0.78), as were Nagelkerke’s R^2^ (0.18 vs 0.28) and scaled Brier’s scores (0.13 vs 0.22). When the outcome was changed to moderately severe asthma (>4 attacks plus use of asthma medication), there were improvements in AUC (0.74), sensitivity (0.97), specificity (0.22) and NPV (0.99), but some deterioration in PPV (0.13), R^2^ (0.15) and scaled Brier score (0.09).

**Conclusion:** While the PARC tool performs well in a population-based setting and has some clinical utility, in particular for ruling out the development of asthma, this study highlights the need for new prognostic prediction tools to be developed specifically for the clinical setting.

**Funding:** SNSF:320030_182628, SLA2019-03_641670

## Introduction

Symptoms such as cough and wheeze in preschool aged children can be an early sign of asthma and cause a high burden of morbidity and health-care utilisation.^1-4^ Asthma also remains an important cause of morbidity later in childhood.^5,6^ To reduce childhood asthma morbidity and overtreatment, it is important for clinicians to identify children with asthma early. For this, they require tools that help distinguish preschool children who are at risk of developing asthma from those who cough or wheeze for other reasons and will possibly outgrow their symptoms.

Several asthma prediction tools have been developed to help identify preschool children at risk of developing asthma. These prediction tools vary in their target population, geography, predictors used and applicable settings. Most use validated questions on respiratory symptoms and some include invasively measured traits such as atopy, fractional exhaled nitric oxide (FeNO), lung function and genetic markers^7-15^.

The PARC tool has the advantage that it predicts asthma risk in preschool children by using questions asked in routine clinical care and avoids the use of invasive measures that may not be available in primary care or resource-poor settings. The tool was developed by Pescatore et. al. using data of preschool children from the population-based Leicester Respiratory Cohort (LRC) who had seen their doctor at age 1-3 years with symptoms of cough or wheeze, and aimed to predict whether they would have asthma 5 years later (Table 1).^16^ The outcome of asthma was defined as wheeze and asthma medication use in the preceding 12 months (Table 1). In internal validation in the LRC, the PARC tool demonstrated good discriminatory performance (area under the curve AUC of 0.78).^16^ In external validation in community-based settings, the PARC tool demonstrated a good discriminatory performance in the Avon Longitudinal Study of Parents and Children (ALSPAC) cohort (area under the curve AUC 0.77)^17^ and very good discriminatory performance in the Multi-centre Allergy Study (MAS) cohort (AUC 0.83)^18^. The PARC tool has yet to be externally validated in clinical settings where preschool children are seen for wheeze or cough. We therefore aimed to perform an external validation of the PARC tool in a prospective cohort of children with respiratory symptoms presenting to paediatric pulmonology outpatient clinics and assess its performance in predicting asthma 2 years later.

**Table 1:** Comparison of Study Characteristics in SPAC versus LRC. SPAC, Swiss Paediatric Airway Cohort; LRC Leicester Respiratory Cohort.

|  | **Development cohort (LRC)** | **External validation cohort (SPAC)** |
| --- | --- | --- |
| Setting | Leicester (United Kingdom) | Aarau, Bern, Basel, Chur, Luzern, St Gallen, Worb, Zurich (Switzerland) |
| Population | 8700 | 3547 |
| Year of birth | 1995-1997 | 2011- 2018 |
| Male sex | 4524 (52%) | 2341 (66%) |
| Study design | Prospective longitudinal cohort | Prospective longitudinal cohort |
| Study population | General population random sample by postal survey circulated in 1990, 1992-94, 1998 and 2003 | Pulmonology clinic-based population recruited at clinic visit since 2017 ongoing on a weekly basis |
| Median age in years at scoring (range) | 2 (1-3) | 3.6 (1-6) |
| Elapsed time between baseline and outcome | 5 years | 2 years |
| Assessment of baseline characteristics | Postal questionnaire | Clinical visit with questionnaire |
| **Inclusion criteria** |  |  |
| Source | Baseline questionnaire from 1998 or 1999 | Baseline questionnaire from 2017 to 2020 |
| Has your child had wheezing or whistling in the past 12 months? | Questionnaire response: Yes, No | Questionnaire response: Yes, No |
| Has your child had cough apart from colds in the past 12 months? | Questionnaire response: Yes, No | Questionnaire response: Yes, No |
| In the last 12 months has your child had a dry cough at night, apart from a cough associated with a cold or a chest infection? | Questionnaire response: Yes, No | Questionnaire response: Yes, No |
| How often did your child see a general practitioner for coughing or wheezing in the last 12 months? | Questionnaire response: Never, once, 2-3 times, 4-6 times, 7 or more times | Child automatically sees doctor as a part of enrolment in SPAC |
| In the last 12 months, has wheezing or asthma resulted in your child: *Being referred to a consultant in hospital; Being admitted to hospital; Attending the casualty (A and E) department; Attending (or calling) the GP in an emergency* | Questionnaire response: Yes, No | Child automatically sees doctor as a part of enrolment in SPAC |
| **Outcome criteria** |  |  |
| Proportion with outcome | 345 (28%) | 125 (36%) |
| Source | Follow up questionnaire 5 years later | Follow up questionnaire 2 years later |
| Has your child had wheezing or whistling in the past 12 months? | Questionnaire response: Yes, No | Questionnaire response: Yes, No |
| Did your child take any of the following drugs during the last 12 months? *A blue inhaler (Salbutamol, Ventolin Bricanyl or other); A brown or orange inhaler (Pulmicort, Flixotide, Becotide, Beclovent or other); A green or green-white inhaler (Serevent or Oxis); A violet or red-white inhaler (Seretide or Symbicort); Steroid tablets (prednisolone) for attacks; Other* | Questionnaire response: Yes, No, Don't know | Questionnaire response: Never, occasionally, less than 3 months, more than 3 months |

## Methods

We used the transparent reporting of a multivariable prediction model for individual prognosis or diagnosis (TRIPOD) guidelines to report this external validation study.^19,20^

### The External Validation Cohort: Swiss Paediatric Airway Cohort (SPAC)

This external validation was performed using data from SPAC, a multi-centre prospective cohort embedded in routine paediatric pulmonology care across Switzerland. SPAC includes children referred for respiratory symptoms such as wheeze, cough and dyspnoea.^21^ As of 28^th^ February 2022, SPAC includes 3547 children aged 0-16 years of whom 1079 have already completed 2 years of follow up (Figure 1). The SPAC study was approved by the Bern Cantonal Ethics Committee (Kantonale Ethikkomission Bern 2016-02176).

**Figure 1:**
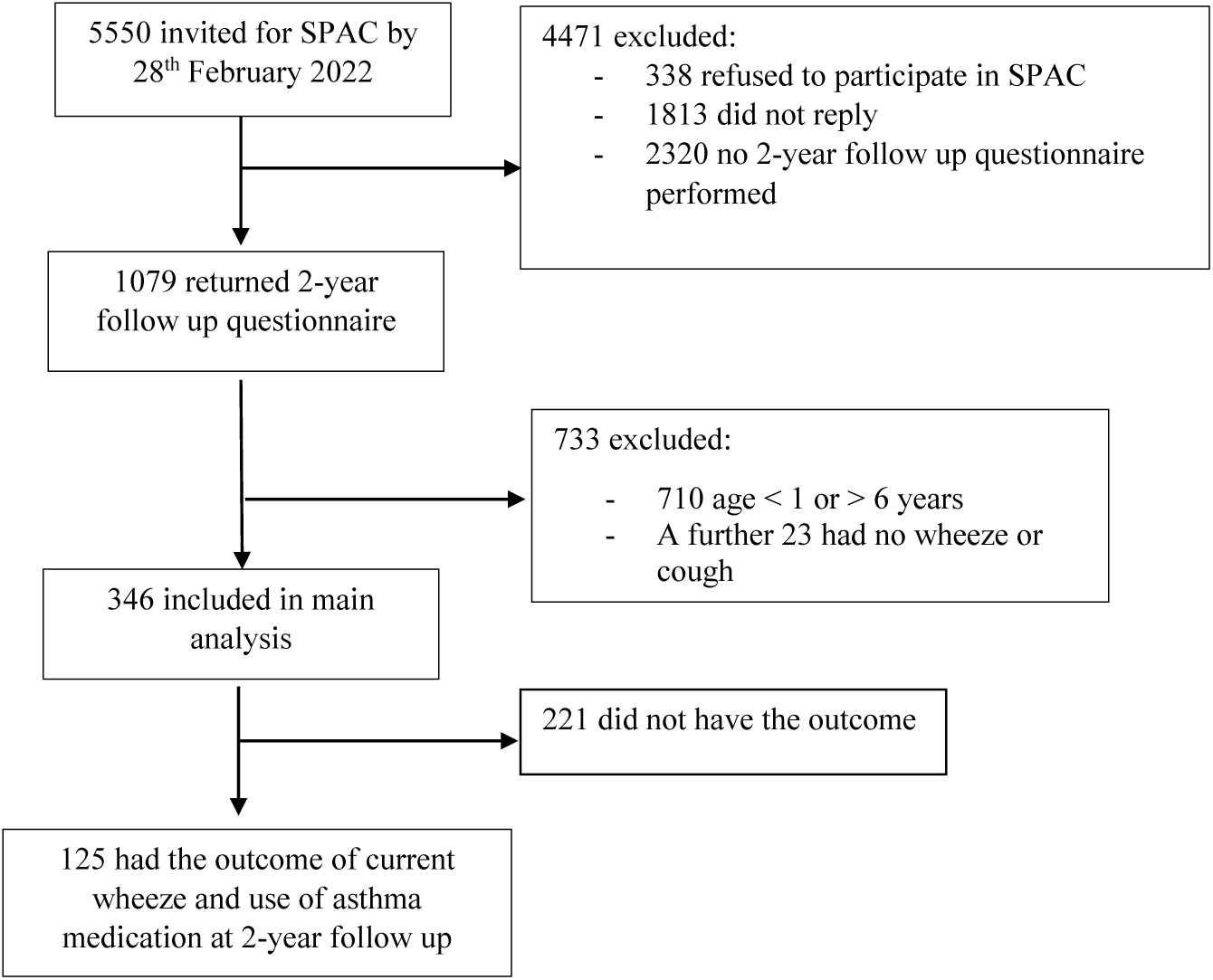
Flowchart of included participants in SPAC. SPAC, Swiss Paediatric Airway Cohort.

### The PARC Tool

The PARC tool includes 10-items in its prediction score. These include: age, sex, occurrence of wheeze episodes not associated with a cold, number of episodes of wheeze, impact of wheezing on daily activities, dyspnoea, wheeze or cough triggered by exercise, laughing, crying or excitement, wheeze or cough triggered by dust, grass, pets or other animals, history of eczema in the child and history of asthma, bronchitis or wheeze in the parents. Each item is assigned a score with the overall sum ranging from 0 to 15.

### Inclusion Criteria

Inclusion criteria for this external validation were selected to resemble original LRC inclusion criteria as closely as possible (Table 2). In order to achieve a larger sample size the inclusion age range was expanded from 1-3 to 1-6 years. Children were included if at the baseline visit they responded “yes” to the question: “In the past 12 months, has your child ever had a dry cough at night, apart from a cough associated with a cold or chest infection?” or “Has your child had wheezing or whistling in the chest in the past 12 months?” and if they had responded to the 2-year follow up questionnaire.

**Table 2:**
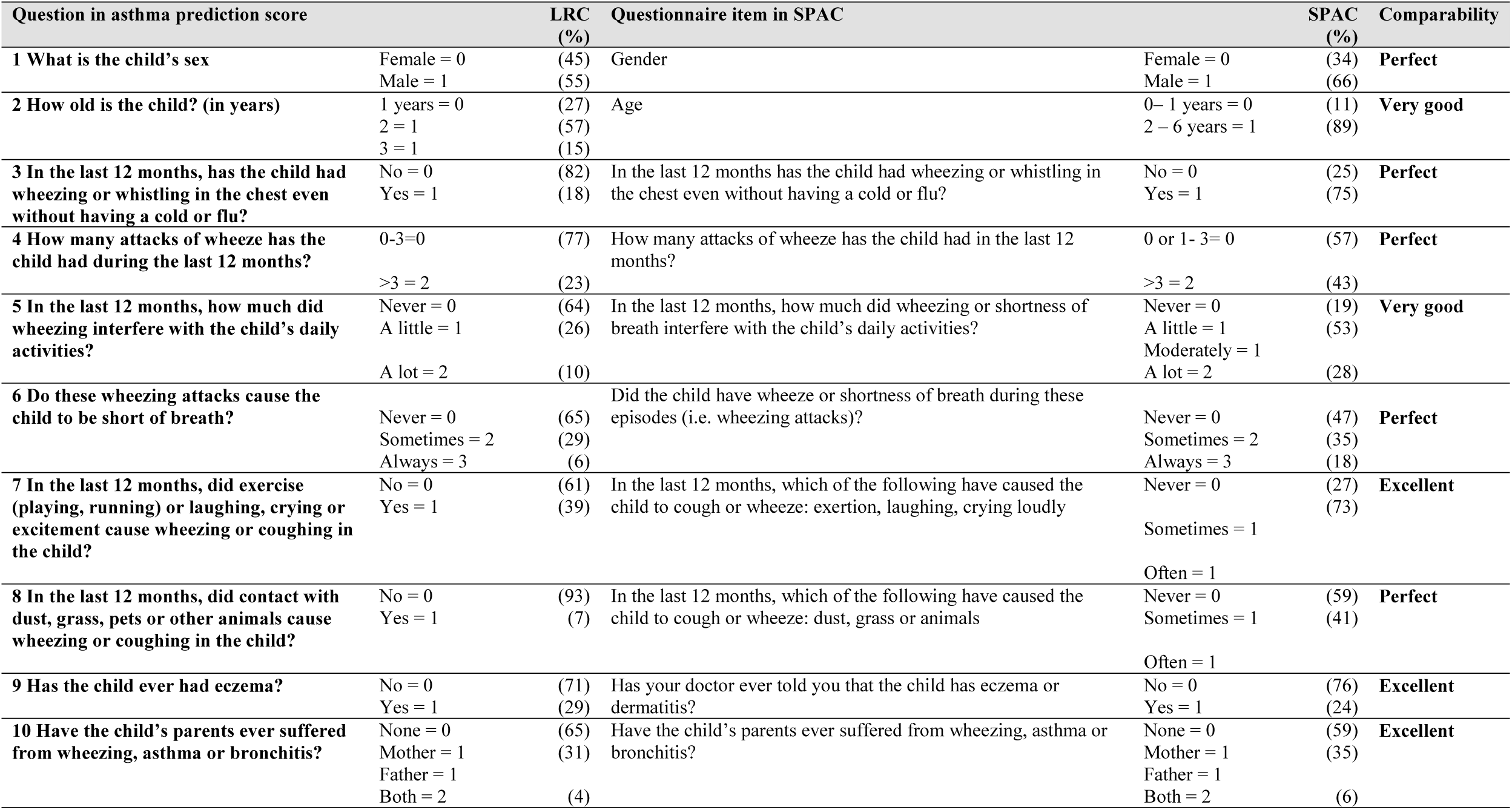
Questions used for each PARC score item at baseline in SPAC and LRC. SPAC, Swiss Paediatric Airway Cohort; LRC Leicester Respiratory Cohort.

### Outcome Criteria

Outcome criteria were selected to resemble original LRC outcome criteria (Table 2). Time to the development of the outcome was shortened from 5 years to 2 years due to current study limitations, as we do not yet have 5 years of follow-up. We defined the outcome as children who responded “yes” to the question: “Has your child reported wheeze in the past 12 months” at the 2-year follow up questionnaire and who also reported having used an asthma medication in the previous 12-months.

### Sample Size

As a rule of thumb, a sample used for external validation should include at least 100 outcome events and 100 non-outcome events.^23^ This external validation is based on 125 events and 221 non-events.

### Missing Data

The data used to assess PARC score predictors was complete for the majority of predictors. For the remainder, the proportion of missing values was <2% for 3 predictors, 9% for 2 predictors, and 29% for 1 predictor. The predictor with the most missing values was derived from the question: “In the last 12 months, which of the following have caused the child to cough or wheeze: dust, grass or animals?” We decided to recode these missing values to “no”, assuming that if the parents had not decisively answered with “yes” the symptom was absent, borderline or mild.

### Statistical Analysis

We assessed predictive performance of the PARC tool in SPAC using measures of discrimination and calibration. We assessed overall performance in terms of “goodness-of-fit” using maximum-scaled Brier’s score and Nagelkerke’s R^2^ scores. For these scores 1 indicates perfect prediction and 0 non-informative prediction. The Brier Score evaluates mean square error of prediction while Nagelkerke’s R^2^ compares likelihoods between the prediction and a non-informative models. We assessed discrimination using sensitivity, specificity, positive predictive value (PPV), negative predictive value (NPV), positive and negative likelihood ratios (LR+, LR-) and AUC. We used an AUC definition of good if > 0.7, very good if > 0.8 and excellent if >0.9.^24^ We assessed calibration by assigning original probabilities derived from the developmental model by Pescatore et. al. of the outcome of asthma to each PARC score. We then recalibrated PARC scores in SPAC by fitting a logistic regression of the outcome on the calculated scores as a linear term and calculating predicted probabilities for each score from the fitted model. We then compared calibration performance of the recalibrated SPAC scores with the original scores. We examined calibration of the PARC tool graphically by plotting the predicted probability for each value of the score against the observed frequency of asthma among children in SPAC with that score value, using the function calibrate.plot and val.prob.ci.2 from the gbm package in R (R Foundation, Vienna, Austria). We used STATA 17 for data preparation, descriptive analysis and logistic regression and RStudio 2021.09.0 to assess model performance.

### Sensitivity Analysis

To further examine the performance of the PARC tool in SPAC we undertook sensitivity analyses using alternative definitions for inclusion and outcome criteria (Table 3). First, we modified inclusion criteria by age group. For outcome sensitivity analysis we modified the timing of the outcome to 1-year and 3-year follow-up so that we could assess the role of different follow up time frames on the performance of the model. As in the development cohort, LRC, and another external validation cohort, ALSPAC, we also did a sensitivity analysis assessing performance for a more severe definition of asthma (Table 3).

**Table 3:**
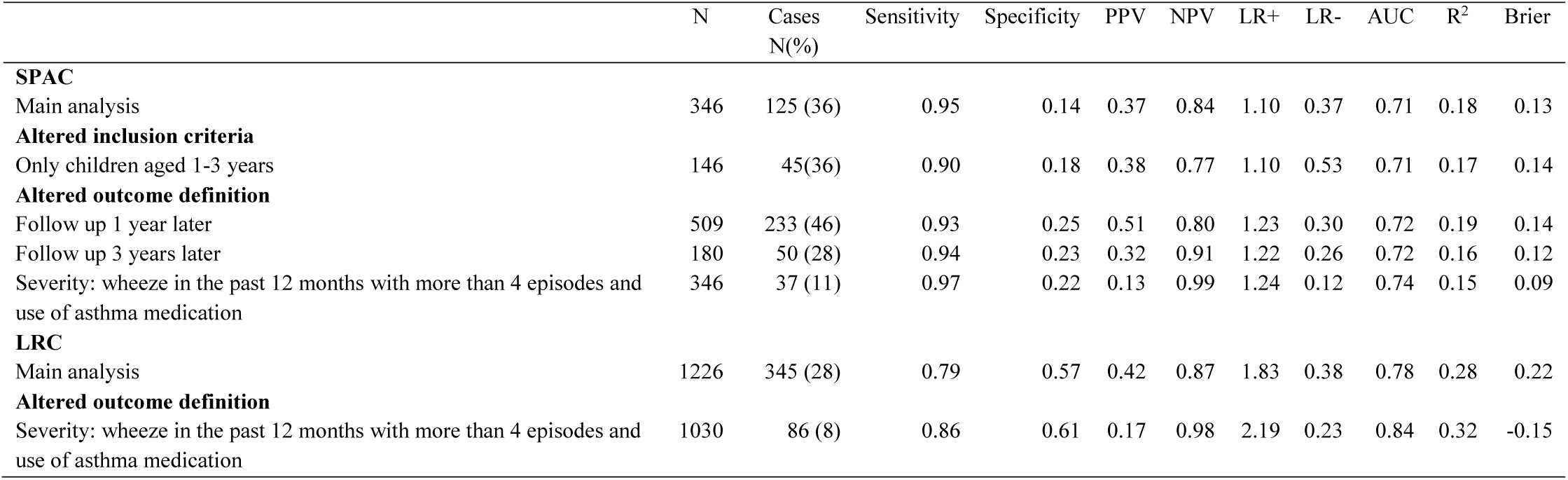
Sensitivity analysis of SPAC versus LRC at a PARC Score of 4

|  | N | Cases<br>N(%) | Sensitivity | Specificity | PPV | NPV | LR+ | LR- | AUC | R<sup>2</sup> | Brier |
| --- | --- | --- | --- | --- | --- | --- | --- | --- | --- | --- | --- |
| **SPAC** |  |  |  |  |  |  |  |  |  |  |  |
| Main analysis | 346 | 125 (36) | 0.95 | 0.14 | 0.37 | 0.84 | 1.10 | 0.37 | 0.71 | 0.18 | 0.13 |
| **Altered inclusion criteria** |  |  |  |  |  |  |  |  |  |  |  |
| Only children aged 1-3 years | 146 | 45(36) | 0.90 | 0.18 | 0.38 | 0.77 | 1.10 | 0.53 | 0.71 | 0.17 | 0.14 |
| **Altered outcome definition** |  |  |  |  |  |  |  |  |  |  |  |
| Follow up 1 year later | 509 | 233 (46) | 0.93 | 0.25 | 0.51 | 0.80 | 1.23 | 0.30 | 0.72 | 0.19 | 0.14 |
| Follow up 3 years later | 180 | 50 (28) | 0.94 | 0.23 | 0.32 | 0.91 | 1.22 | 0.26 | 0.72 | 0.16 | 0.12 |
| Severity: wheeze in the past 12 months with more than 4 episodes and use of asthma medication | 346 | 37 (11) | 0.97 | 0.22 | 0.13 | 0.99 | 1.24 | 0.12 | 0.74 | 0.15 | 0.09 |
| **LRC** |  |  |  |  |  |  |  |  |  |  |  |
| Main analysis | 1226 | 345 (28) | 0.79 | 0.57 | 0.42 | 0.87 | 1.83 | 0.38 | 0.78 | 0.28 | 0.22 |
| **Altered outcome definition** |  |  |  |  |  |  |  |  |  |  |  |
| Severity: wheeze in the past 12 months with more than 4 episodes and use of asthma medication | 1030 | 86 (8) | 0.86 | 0.61 | 0.17 | 0.98 | 2.19 | 0.23 | 0.84 | 0.32 | -0.15 |

## Results

In the SPAC study, 346 of 1079 children (32%) satisfied the inclusion criteria (response yes to wheeze or cough in the past 12 months at baseline), were aged 1-6 years and completed the 2-year follow up. 125 (36% of the 346 children) had the defined outcome. In contrast, in LRC 1226 of 8700 children (14%) satisfied inclusion criteria and 345 (28%) had the outcome. Notable differences between the cohorts include: geographical area, study years (2017-2020 versus 1998-1999) and study setting (clinical cohort versus population-based cohort) (Table 1). Follow up time was 2 years in SPAC and 5 years in LRC. ^22^

The median PARC score was higher in SPAC (median 8, inter-quartile range (IQR) 5 – 8) than in LRC (median 4, IQR 2-6) (Figure 2). At a PARC score of 4, sensitivity was higher (95% versus 79%) and specificity lower (14% versus 57%) in SPAC compared to LRC. The positive and negative predictive values were comparable (37% versus 42%) and (84% versus 87%) in SPAC and LRC respectively. The positive and negative likelihood ratios were also comparable (1.1 versus 1.83 and 0.38 versus 0.38) in SPAC and LRC respectively (Table 3). The PARC tool’s ability to discriminate between those children with and without the outcome was modest in SPAC (AUC 0.71) and lower than in LRC (AUC 0.78) (Figure 3). The tool also showed poorer overall performance in SPAC than LRC with scaled Brier’s score 0.13 versus 0.22 and Nagelkerke’s R^2^ 0.18 versus 0.28 (Table 3).

**Figure 2:**
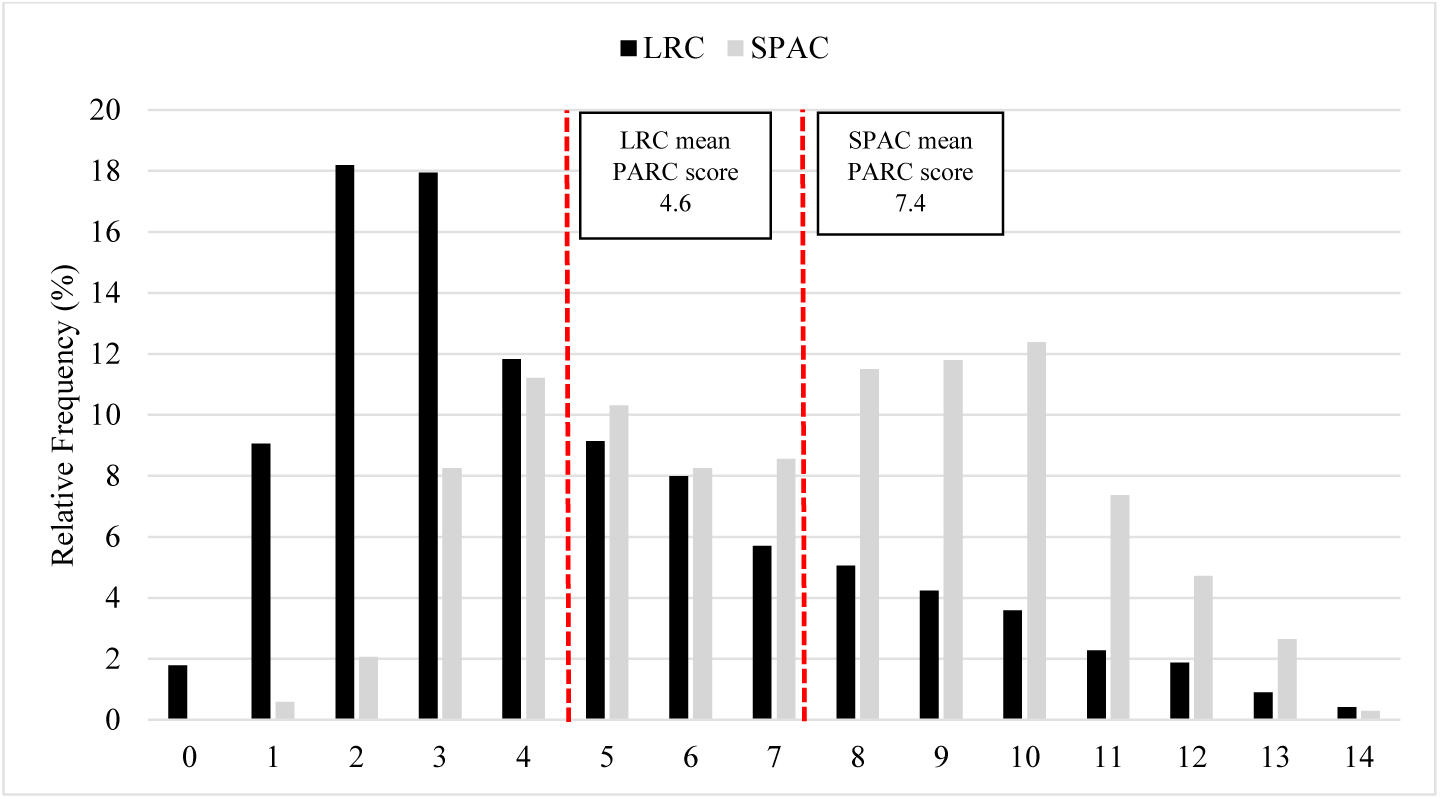
Distribution (relative frequency %) of PARC Score in the external validation population (Swiss Paediatric Airway Cohort, SPAC, N = 346) versus original development population (Leicester Respiratory Cohort, LRC, N = 1226). Mean PARC scores of 7.4 for SPAC and 4.6 for LRC indicated by dashed vertical red lines.

**Figure 3:**
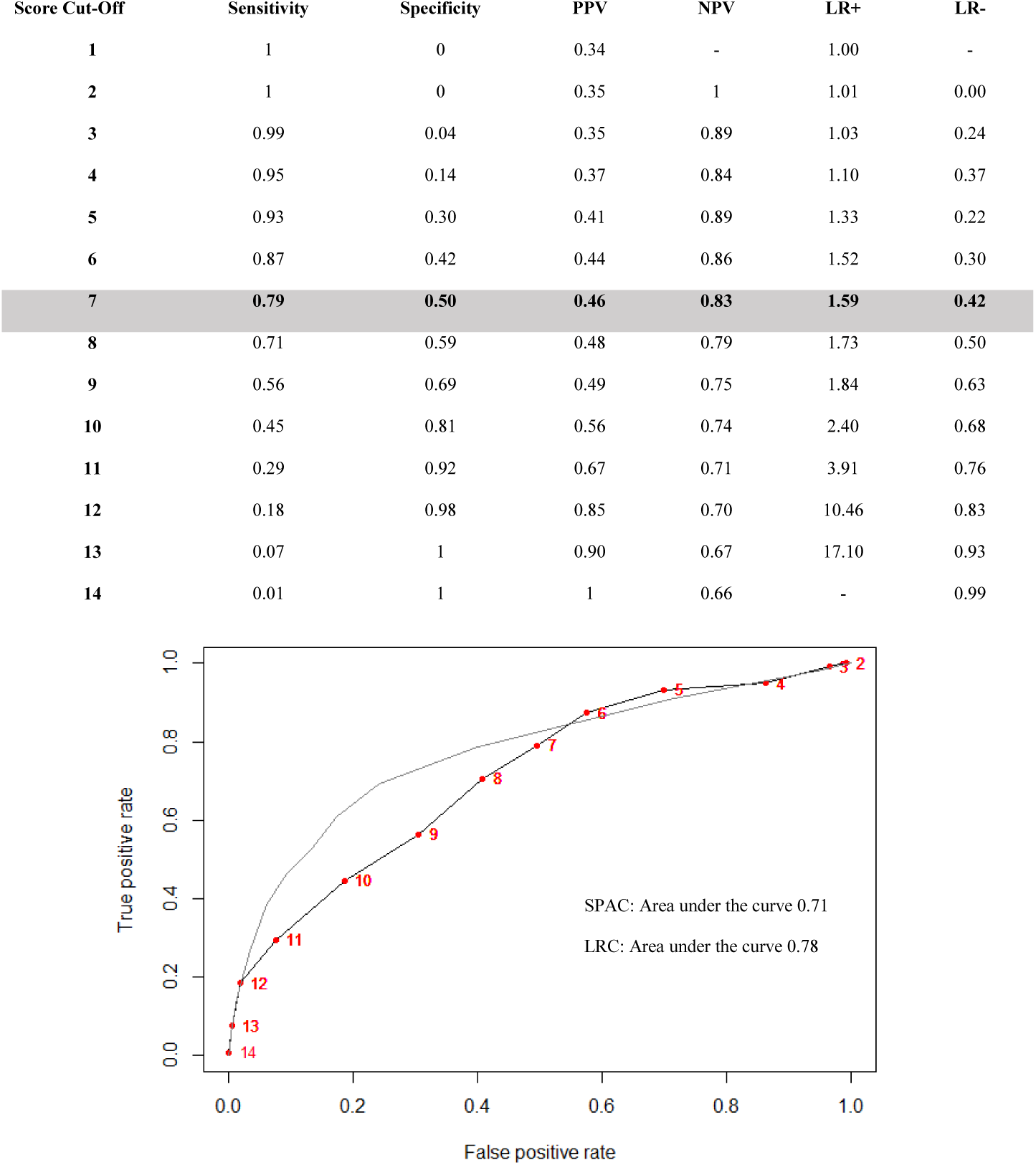
Receiver operating characteristic from external validation (SPAC in black with PARC scores marked in red) and development cohort (LRC in grey) including area under the curve. Table shows sensitivity, specificity, positive predictive value, negative predictive value, positive likelihood ratio and negative likelihood ratio for each score in SPAC. SPAC, Swiss Paediatric Airway Cohort; LRC, Leicester Respiratory Cohort.

Sensitivity analysis with changes to inclusion and outcome criteria in SPAC resulted in modest adjustments to performance indicators (Table 3). Altering inclusion criteria to 1-3 year-olds only as per LRC inclusion criteria resulted in AUC of 0.71. Altering the outcome criteria to a higher severity defined as > 4 episodes of wheeze in the past 12 months plus asthma medication use improved the AUC to 0.74. This same sensitivity analysis resulted in an AUC of 0.84 when performed in the LRC.^17^ Altering outcome criteria to follow up 1 year or 3 years later increased the AUC marginally to 0.72.

Assessment of performance at a PARC score of 7 showed higher sensitivity (79% versus 46%), lower specificity (51% versus 91%), comparable PPV and NPV (45% versus 44% and 83% versus 91%), lower positive likelihood ratio (1.6 versus 4.95) and comparable negative likelihood ratio (0.42 versus 0.59) in SPAC versus LRC (Figure 2). The maximum predicted probability of developing the outcome in SPAC was 76% compared to 95% in LRC (Figure 3). The Loess smoothed calibration plot demonstrated c statistic of 0.71 (CI 0.65 to 0.77), slope of 1 (CI 0.69 - 1.31) and intercept of 0 (−0.24 to 0.24) (Figure 4).

**Figure 4:**
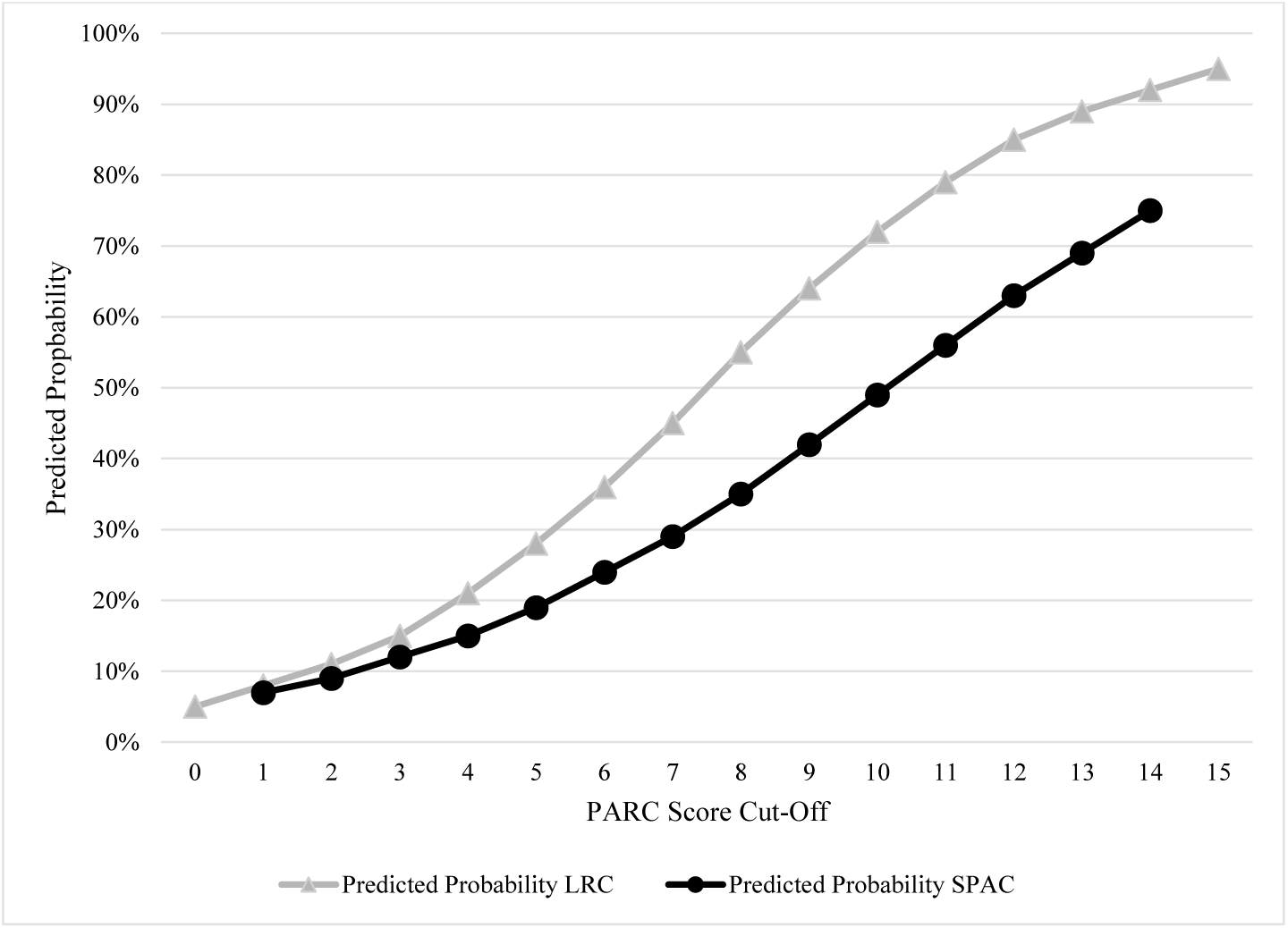
Predicted probability of developing asthma at follow-up in the LRC (grey line) and probability of developing asthma in recalibrated model in SPAC (black line). SPAC, Swiss Paediatric Airway Cohort; LRC Leicester Respiratory Cohort.

**Figure 5:**
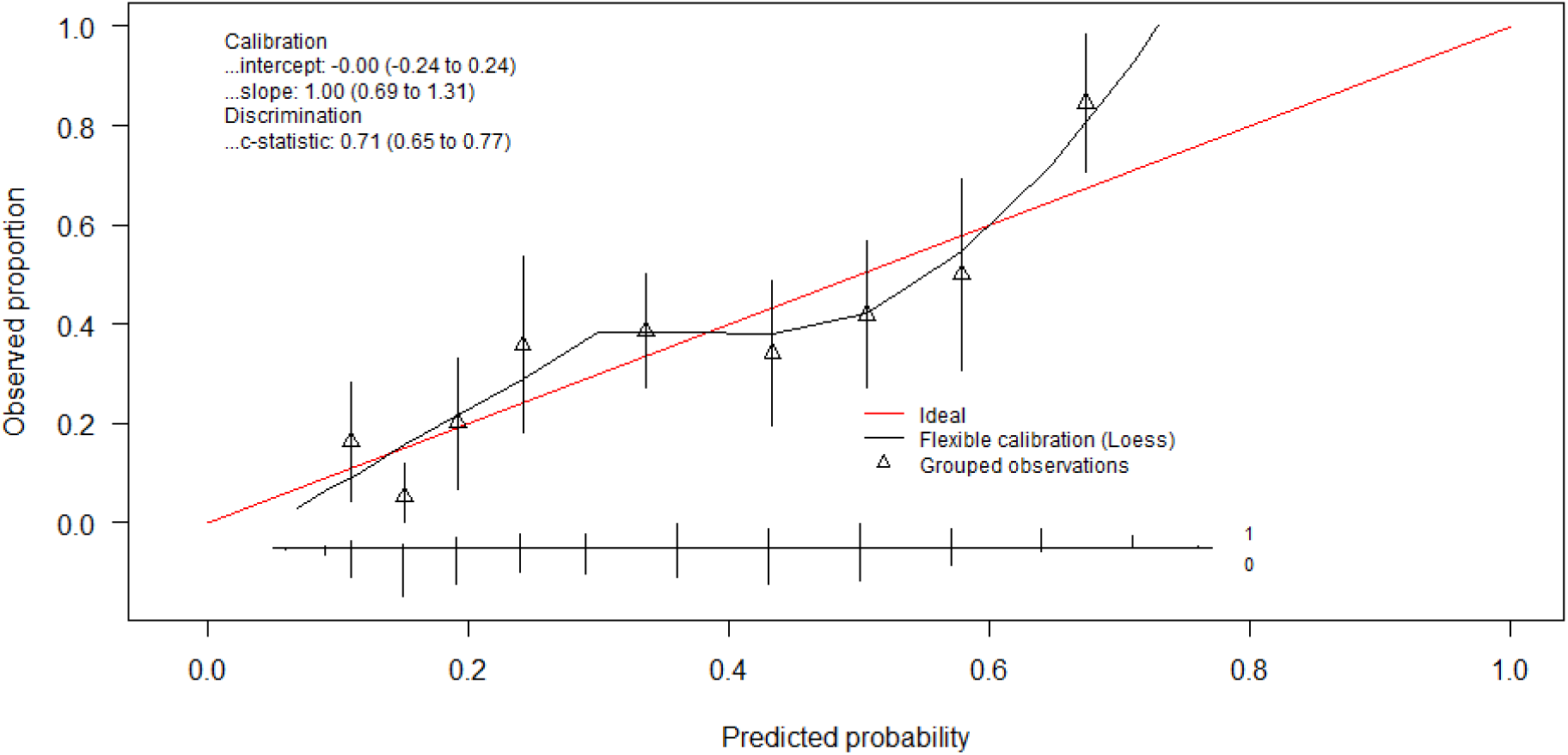
Calibration assessment of predicted probabilities in SPAC. SPAC, Swiss Paediatric Airway Cohort; LRC Leicester Respiratory Cohort.

## Discussion

This is the first external validation of the PARC tool in a clinical cohort. The tool showed reduced predictive ability for asthma development in young children aged 1-6 years old in the clinical validation cohort (SPAC) than in the original population-based cohort (LRC). Discrimination, though modest in both cohorts, was somewhat reduced in SPAC (AUC 0.71) compared to LRC (0.78). The PARC tool had a higher sensitivity (95% versus 79%) at a score of 4 in the clinical setting of SPAC compared to the community-setting of LRC, and a similarly high negative predictive value (84% vs 87%). However, all other indicators suggested reduced performance. The mean PARC score was higher in SPAC than LRC and AUC improved to 0.74 when the outcome was defined as moderately severe asthma (more than 4 attacks in the past year and use of asthma medication).

### Strengths and limitations

This study has several strengths. It is the first external validation of the tool in a clinical setting where children with respiratory problems are treated. In addition, SPAC is a longitudinal study embedded in routine pulmonology care across German-speaking parts of Switzerland, which represents a real-world setting. Questionnaire items used in SPAC are identical to those used in LRC. A strength of the PARC tool is, that it used state-of the art modelling strategies and selected predictors using a method (least absolute shrinkage and selection operator (LASSO) regression) that minimises the risk of over-fitting the data. The main limitation of this external validation is the small sample size currently available in this clinical setting and the limited follow-up time. However, due to the longitudinal and ongoing nature of SPAC, there is scope to perform further external validations in future when the participant number and follow up time has increased.

### Interpretation and Comparison with Previous Studies

There are several differences between SPAC and LRC which must be considered when interpreting the performance of the PARC tool in this external validation setting. Firstly, the broader age range (1-6 years versus 1-3 years) and shorter follow up time (2 years versus 5 years) in SPAC compared to LRC. Secondly, this analysis used information collected from parent-reported questionnaires written in German. The specific terms for “wheeze” are not easily interchangeable between the German and English languages. However, a previous external validation of the PARC tool in the German MAS population-based cohort showed very good discriminatory performance with an AUC of 0.83, suggesting that differences in language are not a likely explanation for the poorer performance of PARC in SPAC.^18^ Thirdly, SPAC represents a group of children with a high burden of respiratory disease in a clinical setting. A higher burden of respiratory disease results in a more homogenous case-mix which has been shown in external validations to reduce discriminative ability of prediction models^25^ and application of a prediction tool from a population or primary care setting to a tertiary setting has also been shown to demonstrate reduced spectrum transportability^26^.

Other asthma predictive tools have been developed for preschool children in clinical settings. A retrospective study by Boersma et. al. that aimed to examine how sensitisation to inhalant allergens among wheezing toddlers in secondary healthcare predicted asthma development, demonstrated an improvement in the AUC from 0.70 to 0.79 when it included invasive sensitisation measures compared to when they used only International Study of Asthma and Allergies in Childhood (ISAAC) questions.^27^ Vial Dupuy et. al developed and independently validated the Persistent Asthma Predicting Score (PAPS) in a clinical setting. They defined the outcome as “persistent” asthma. Predictors included: family history of asthma, personal history of atopic dermatitis and IgE sensitisation. Their analysis demonstrated an AUC of 0.66 in the development population and 0.65 in the internal validation population demonstrating low overall discrimination of the tool.^14^ In a clinical setting including corticosteroid-naïve 3-47 month old children in Zürich, Switzerland, Singer et. al. performed an external validation of a modified Asthma Predictive Index (API) tool. They used an outcome of physician diagnosed asthma and included invasively-measured FeNO as a predictor. Their analysis showed good discrimination with AUC of 0.76 when FeNO was used instead of blood eosinophilia, as per the original API tool.^28^ Rodriguez-Martinez et. al. implemented the loose API and modified Prevalence and Incidence of Asthma and Mite Allergy (PIAMA) tools in a clinical setting in Bogota, Columbia. They reported a sensitivity and specificity for predicting asthma at age 5-6 years in pre-schoolers with recurrent wheeze of 71.4% and 33.3% (loose API) and 54.5% and 78.9% (modified PIAMA) for ideal ROC cut off points. The authors suggested that these prediction tools could therefore be applicable in clinical practice.^29^ In our external validation, the PARC tool had an NPV of 83%, PPV of 46%, and sensitivity and specificity of 79% and 50% respectively at a PARC score of 7, which suggests that it also has scope for clinical application.

### Implications for Clinical Care and Future Research

The PARC tool has now undergone external validation in two population-based and one clinical cohort. The tool shows good to very-good discriminative ability in community settings and good discriminative ability in the clinical setting. However, it has a higher sensitivity of 95% and an NPV of 84% in SPAC, indicating a possible role for assisting clinicians to accurately identify children with a low risk of developing. For example, paediatric pulmonologists in Switzerland could use the PARC tool to reassure parents with children with a score of 4 or less that their child’s risk of having asthma in 2 years’ time is only 5%. The proportion of children in our analysis with a PARC score 4 or less was 10%. The tool could also have potential applications for triaging patients for follow up. Furthermore, by narrowing the definition of asthma to a more severe spectrum the tool performs better in SPAC. This suggests that, in a clinical context, the PARC tool may be able predict severe asthma better and that any adaptation of the PARC tool in the future should use different outcome definitions such as “severe asthma” or “asthma persistence”. Previous applications of prediction tools to clinical settings have demonstrated a role for invasive measures such as blood eosinophils, IgE sensitisation and FeNO with variable success.^14, 26, 27^ Future prediction model development for asthma risk in preschool children attending respiratory clinics could include these invasive measures to improve their discriminatory performance.

## Conclusion

This external validation of the PARC tool in a clinical cohort embedded in routine paediatric pulmonology care suggested that the tool has a role for application in this setting. Currently, it can be used to help identify 1-6 year old children who are at low risk of having asthma at 2-years of follow up, due to its negative predictive value and sensitivity at a cut-off of 4. Discrimination of the PARC tool between children who will and will not have asthma 2 years later was borderline good highlighting the need for new childhood asthma prediction tools in the clinical setting. We suggest that future efforts to improve asthma prediction in this setting should explore the added value of biomarkers of physiological asthma-related traits available in pulmonology clinics, such as FeNO, allergy tests and lung function measurements.

## Supporting information

TRIPOD Checklist

## Data Availability

All data produced in the present study are available upon reasonable request to the authors.

## Acknowledgements

Members of the SPAC Study Team are: D. Mueller-Suter and P. Eng (Canton Hospital Aarau, Aarau, Switzerland); U. Frey, J. Hammer, A. Jochmann, D. Trachsel, and A. Oettlin (University Children’s Hospital Basel, Basel, Switzerland); P. Latzin, C. Abbas, M. Bullo, C. Casaulta, C. de Jong, E. Kieninger, I. Korten, L. Krüger, F. Singer, and S. Yammine (University Children’s Hospital Bern, Bern, Switzerland); P. Iseli (Children’s Hospital Chur, Chur, Switzerland); K. Hoyler (private paediatric pulmonologist, Horgen, Switzerland); S. Blanchon, S. Guerin, and I. Rochat (University Children’s Hospital Lausanne, Lausanne, Switzerland); N. Regamey, M. Lurà, M. Hitzler, K. Hrup, and J. Stritt (Canton Hospital Lucerne, Lucerne, Switzerland); J. Barben (Children’s Hospital St Gallen, St Gallen, Switzerland); O. Sutter (private paediatric practice, Worb, Bern, Switzerland); A. Moeller, A. Hector, K. Heschl, A. Jung, T. Schürmann, L. Thanikkel, and J. Usemann (University Children’s Hospital Zurich, Zurich, Switzerland); and C.E. Kuehni, C. Ardura-Garcia, D. Berger, M.C. Mallet, E. Pedersen, and M. Goutaki.

## Abbreviations

ALSPAC: Avon Longitudinal Study of Parents and Children
API: Asthma Predictive Index
AUC: Area under the curve
FeNO: Fractional exhaled nitric oxide
IgE: Immunoglobulin E
IoW: Isle of Wight
IQR: inter-quartile range
ISAAC: International Study of Asthma and Allergies in Childhood
LASSO: least absolute shrinkage and selection operator
LR: Likelihood Ratio
LRC: Leicester Respiratory Cohort
MAS: Multi-centre Allergy Study
PAPS: Persistent Asthma Predicting Score
PARC: Predicting Asthma Risk in Children
PIAMA: Prevalence and Incidence of Asthma and Mite Allergy
ROC: Receiver operator curves
SPAC: Swiss Paediatric Airway Cohort

## Notes

### Competing Interest Statement

The authors have declared no competing interest.

### Author Declarations

The ethics committee of the University of Bern gave ethical approval for this work (Kantonale Ethikkomission Bern 2016-02176).

